# *Coccidioides* genomes from low-incidence states reveal complex migration history across the Western United States

**DOI:** 10.1101/2025.05.19.25327631

**Authors:** Emanuel M. Fonseca, Shanaya Fox, Adrienne L. Carey, Bridget Barker, Marco Marchetti, Megan Hirschi, Kimberly E. Hanson, Katharine S. Walter

**Author notes:** **Corresponding author:** Emanuel M. Fonseca.

## Abstract

The evolutionary history of *Coccidioides immitis* and *C. posadasii*, the fungal agents of coccidioidomycosis or Valley fever, can provide insights into historic and ongoing patterns of dispersal. However, in regions with low Valley fever incidence, genomic data are limited. To better characterize *Coccidioides* diversity and evolutionary history in low incidence states, we prospectively collected *Coccidioides*-positive isolates submitted to a national diagnostic laboratory between January 2023 and November 2024. We conducted whole-genome sequencing for 27 isolates from Utah, Colorado, and Nevada— states with limited genomic characterization. Additionally, we sequenced 22 isolates from California. We assessed geographic structure with maximum likelihood phylogenies, including newly sequenced and previously sequenced *Coccidioides* isolates, and inferred historic patterns of dispersal with ancestral area reconstruction. For patients within the University of Utah Health system, we conducted medical chart reviews to collect information about potential travel, occupational, and recreational exposure history. Among the newly sequenced isolates, 3/27 were C. immitis and 24/27 *C. posadasii*. We identified both C. immitis (3 isolates from 1 individual patient) and *C. posadasii* (16 isolates from 10 individual patients) in Utah. The individual infected with C. immitis reported a history of travel to southern California, consistent with a travel-associated infection. Only *C. posadasii* was detected in Colorado and Nevada. Newly sequenced isolates from Utah, Nevada, and Colorado did not fall into state-specific monophyletic clades, but were instead distributed across the *C. posadasii* phylogeny. Our collection of C. immitis likely originated in California (likelihood = 0.96), while *C. posadasii* traces its ancestral area to Arizona (likelihood = 0.99). We inferred at least seven independent introductions of *C. posadasii* into Utah, two into Nevada, and seven into Colorado. Our findings suggest a complex dispersal history of *Coccidioides* across the western U.S., with evidence of ongoing and repeated dispersal rather than a single historical emergence event. These findings highlight the importance of continued genomic surveillance to better understand the evolutionary history of *Coccidioides* in regions outside highly endemic areas.

## 1. Introduction

*Coccidioides immitis* and **C. posadasii** are soil-dwelling fungal pathogens that cause coccidioidomycosis, commonly known as Valley fever. This disease, which ranges from mild respiratory illness to severe systemic infection, results from the inhalation of airborne arthroconidia produced by these fungi (Brown et al., 2013; Galgiani et al., 2016). Endemic to arid and semi-arid regions of the Americas, the two species exhibit distinct geographic distributions. *Coccidioides immitis* is primarily found in California and Baja Mexico, though recent evidence suggests its range is expanding, with newly identified isolates in Washington and Oregon (Kirkland & Fierer, 2018; Monroy-Nieto et al., 2023). In contrast, **C. posadasii** spans a broader area, including the southwestern United States (Arizona, Texas, and New Mexico), Mexico, and parts of Central and South America (Kirkland & Fierer, 2018; Monroy-Nieto et al., 2023).The ecological niche of these fungi is closely linked to specific environmental conditions—warm temperatures, seasonal rainfall followed by dry periods, and elevated dust levels—that promote their growth and dispersal (Comrie, 2005; Gorris et al., 2019).

Coccidioidomycosis is increasingly recognized as a re-emerging infectious disease, with rising incidence rates in endemic regions and an expanding geographic footprint (Camponuri et al., 2024; Johnson et al., 2014; Marsden-Haug et al., 2013). Recent modeling work has found that the expansion of reported cases of coccidioidomycosis is driven by climate change and land use alterations (Gorris et al., 2019; Head et al., 2022). For instance, warmer temperatures and prolonged droughts may enhance fungal growth, while urbanization and agricultural activities disturb soil, releasing spores into the air.

Despite its growing public health significance, coccidioidomycosis remains underdiagnosed and underreported due to its nonspecific symptoms and limited awareness among some healthcare providers (Benedict et al., 2021; Gorris et al., 2023). As the distribution of *Coccidioides* continues to extend beyond historically endemic areas, there is an urgent need to better understand its evolutionary history and historic migration patterns of *Coccidioides* to understand ongoing and future dispersal.

While extensive genomic research has focused on high-incidence endemic regions (e.g., Monroy-Nieto et al., 2023; Neafsey et al., 2010; Oltean et al., 2019), low-incidence states such as Utah, Colorado, and Nevada remain poorly characterized in terms of genomics, despite documented cases of coccidioidomycosis. The lack of genomic data from these regions means that which *Coccidioides* species is locally endemic is not confirmed. For example, one study reported that both species were present at a single sampling site in Dinosaur National Park, in northeastern Utah (Johnson et al., 2014).

Further, it limits our ability to reconstruct migration patterns and identify potential geographic reservoirs for the fungus, responsible for seeding introductions elsewhere.

Here, we characterized the evolutionary history of *Coccidioides* in low-incidence states. We collected and sequenced clinical isolates from Utah, Colorado, and Nevada and, for isolates from Utah, collected detailed travel and exposure history. We combined a *Coccidioides* phylogeny with information on location of Valley fever patients to reconstruct historic patterns of *Coccidioides* spread.

## 2. Results

### 2.1. Species Distribution Across Endemic and Low-Incidence States

Among the 27 newly sequenced isolates from low-incidence states (Utah, Colorado, and Nevada) (Fig 1), 24 were identified as **C. posadasii**, while only three were *C. immitis*.

**Fig 1.**
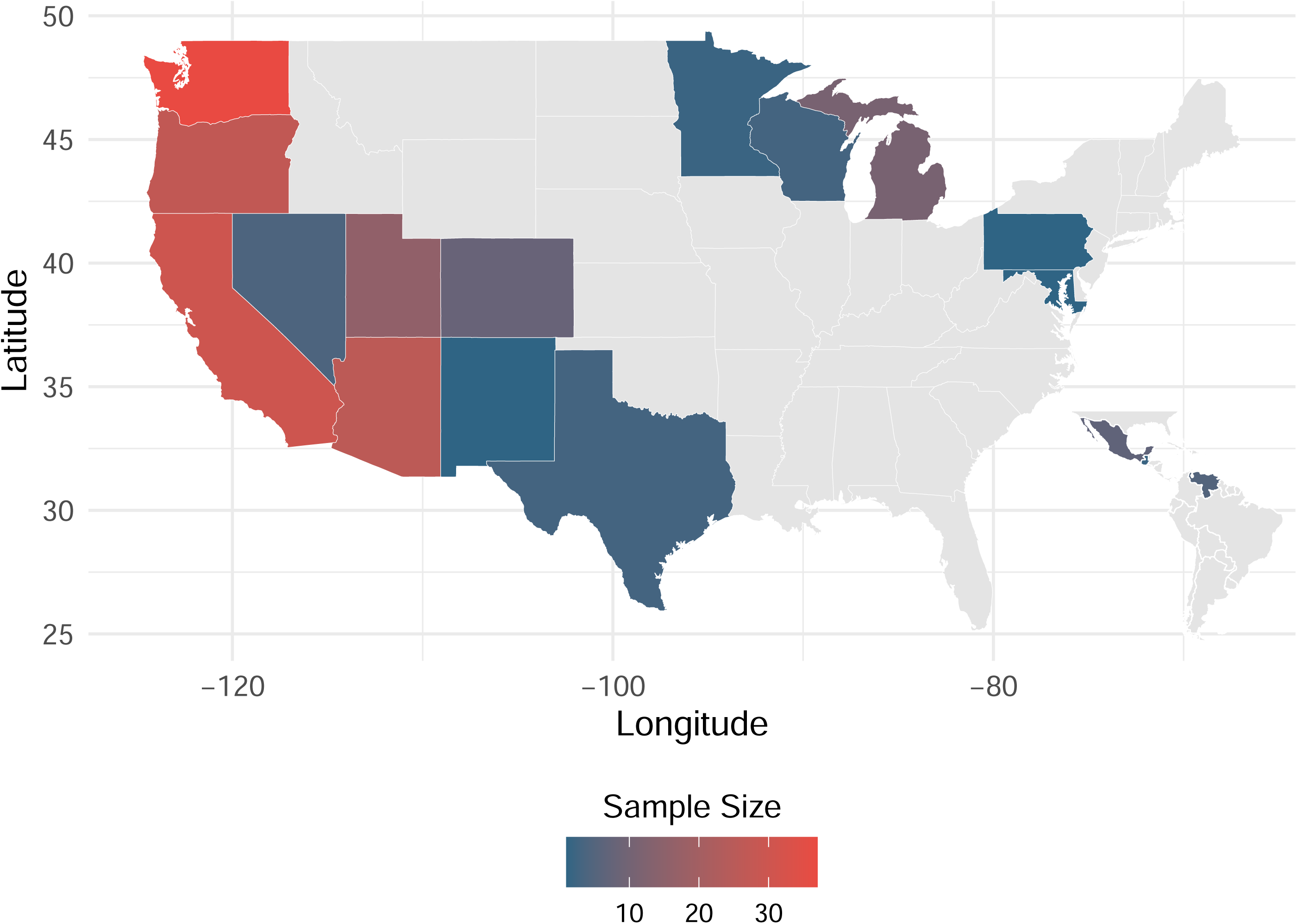
Sample sizes of sequenced *Coccidioides* genomes from across U.S., represented by a gradient color scale from blue (smallest sample size) to red (largest sample size). The inset map highlights samples collected outside the United States.

Notably, Utah was the only low-endemicity state where both species co-occurred, with *C. immitis* detected in 3 isolates from a single patient and **C. posadasii** found in nine (Fig 2). In contrast, **C. posadasii** was the sole species detected in Colorado and Nevada (Fig 2). In high-endemicity states, *C. immitis* was predominant in California, where 20 out 22 newly sequenced isolates belonged to this species. In contrast, **C. posadasii** was more prevalent in Arizona, as reflected by the broader sample representation in this region, including the two remaining sequenced isolates from California (Fig 2).

**Fig 2.**
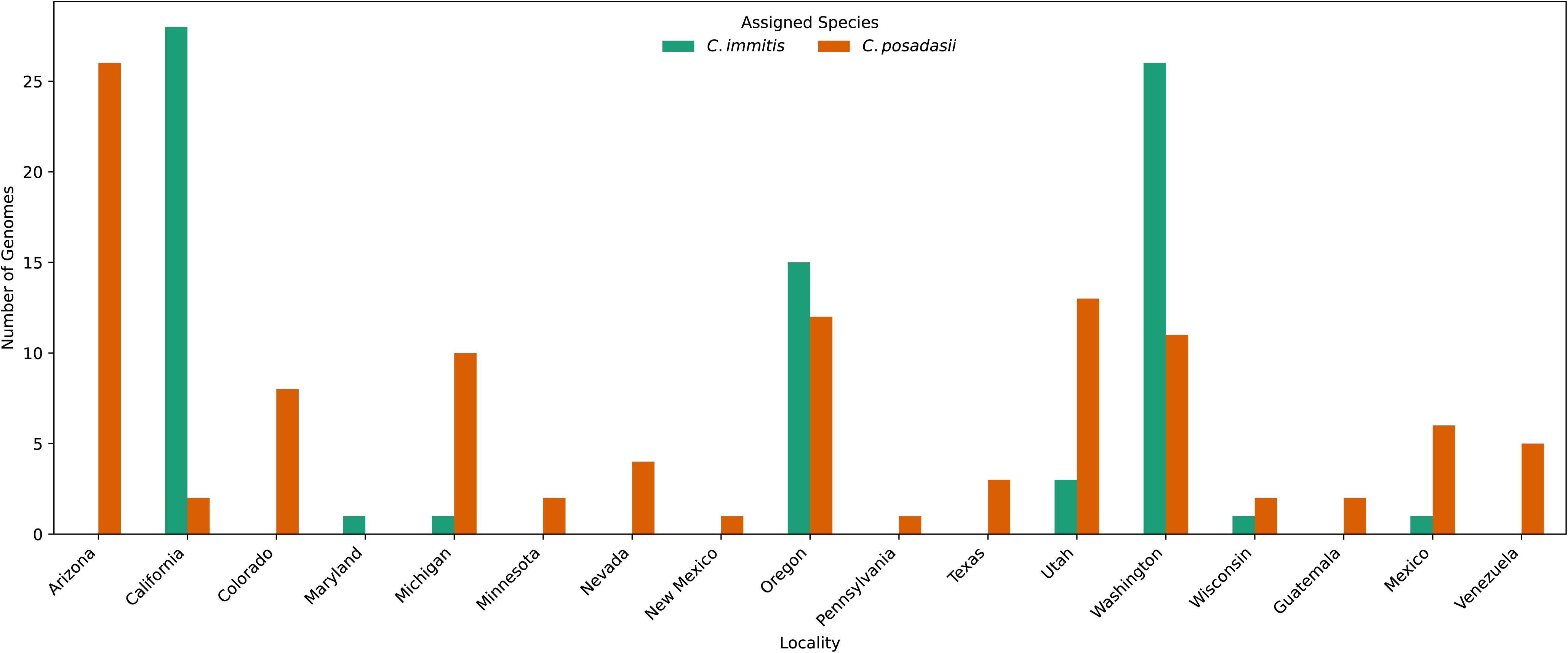
Species identification across sampled localities. Bars represent the number of samples per locality, with *Coccidioides immitis* shown in blue and **C. posadasii** in orange. The x-axis denotes sampled localities, while the y-axis indicates the corresponding sample count.

### 2.2. Epidemiological Profiles and Exposure Histories of Utah Patients

Only four of the ten patients participated in our chart review. These four patients from Utah, who completed our questionnaire, ranged in age from 41 to 70 years and included three women and one man, with ages distributed across the 40–50, 50–60, and 60–70 year age ranges. Among them, only one individual reported a travel history to San Diego and Alaska and had previously lived in southern California, while the others had no recorded travel history. This individual was infected with *C. immitis* and was likely exposed outside of Utah. In contrast, the 10 other patients were infected with **C. posadasii**, suggesting that this species is the only endemic species in Utah.

### 2.3. Phylogenetic Evidence for Multiple Independent Introductions

In a maximum likelihood phylogeny of 185 *Coccidioides* samples, isolates from Utah, Colorado, and Nevada did not form monophyletic state-specific clades but were instead distributed across multiple lineages (Fig 3a–b). This pattern suggests multiple independent introductions into low-endemicity states rather than a single expansion event (Fig 3a). Phylogenetic analysis supports this observation, as geographically proximate isolates did not group strictly according to state boundaries. Additionally, the single Utah isolate of *C. immitis*, obtained from one patient, likely represents an infection acquired through travel, as the patient had documented travel history to San Diego and previously resided in Southern California.

**Fig 3.**
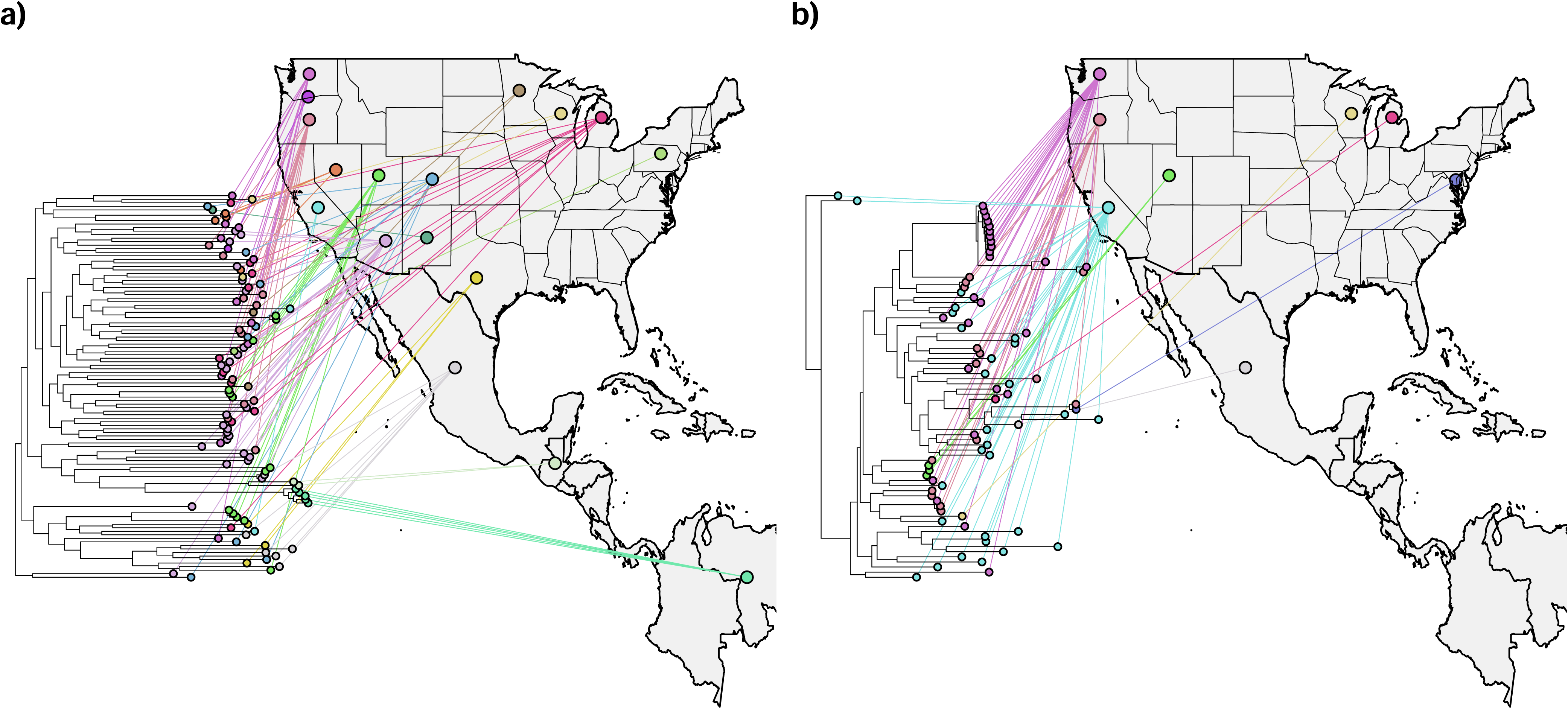
Maximum likelihood phylogenies illustrating the evolutionary relationships among sampled isolates of *Coccidioides*, with panel (a) showing **C. posadasii** and panel (b) showing *C. immitis*. The accompanying maps indicate the U.S. state and country of origin for each isolate, highlighting patterns of geographic distribution and phylogenetic clustering.

### 2.4. Inference of Geographic Origins and Dispersal Pathways Using Ancestral Area Reconstruction

Ancestral area reconstruction revealed multiple independent introductions of **C. posadasii** into Utah, Nevada, and Colorado (Fig 4). Utah received at least seven introductions, primarily originating from Arizona (estimated probability: 50–60%), with secondary contributions from Texas (20–30%) and California (10–20%). Nevada experienced two introductions, most likely from California (70–80%), with a smaller contribution from Arizona (20–30%). Colorado also had seven introductions, predominantly sourced from Arizona (40–50%) and Texas (30–40%), with additional input from New Mexico (10– 20%). Minor introductions from other endemic regions, such as Mexico or Utah, were detected but occurred with lower probability (probabilities <10%). These patterns highlight the role of neighboring endemic areas, particularly Arizona, Texas, and California, in seeding **C. posadasii** populations in low-incidence states.

**Fig 4.**
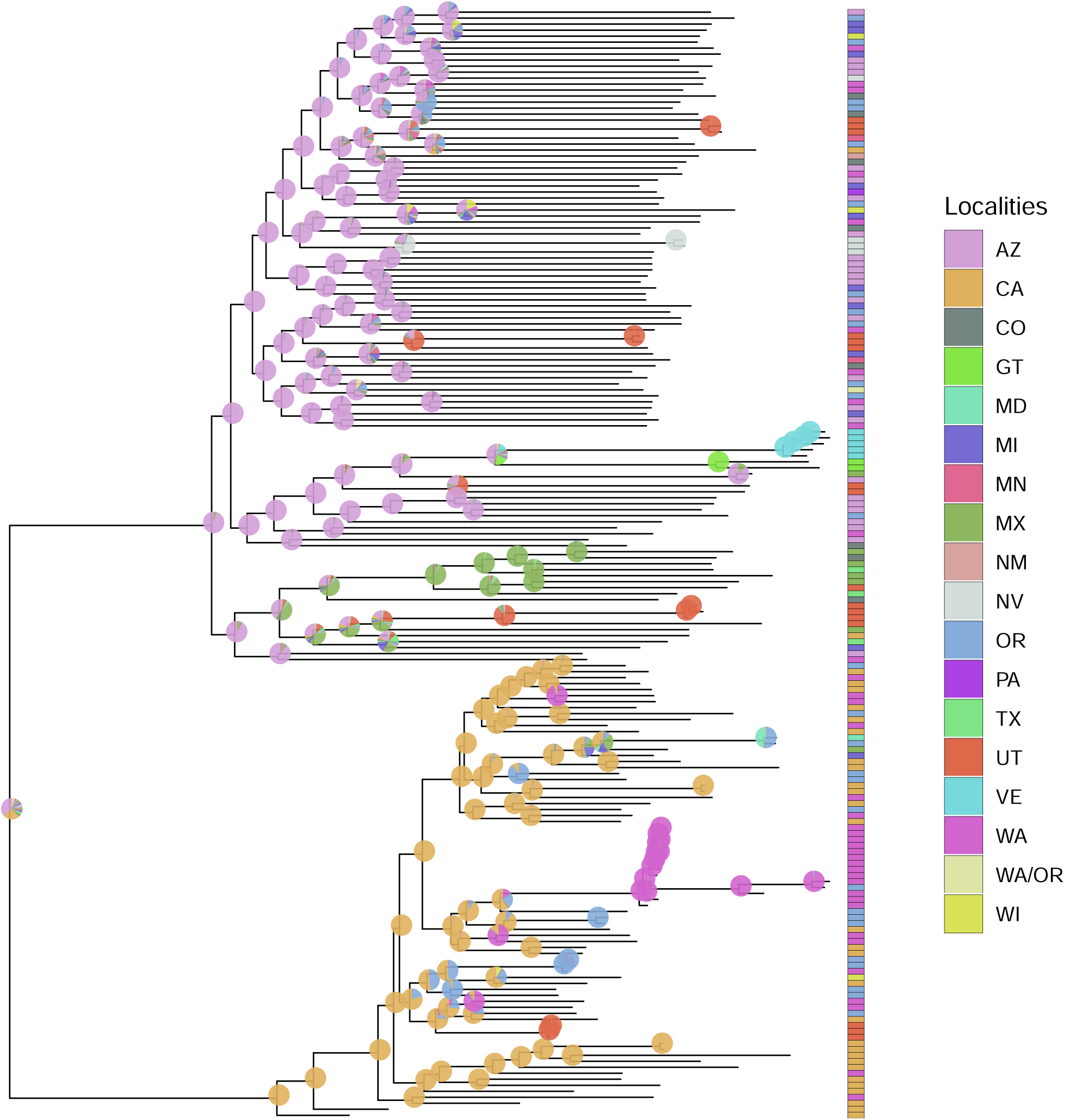
Maximum likelihood phylogeny illustrating ancestral area reconstruction across sampled isolates. Pie graphs on internal nodes represent the probability the ancestor occurred at a specific location, with possible localities including Arizona (AZ), California (CA), Colorado (CO), Guatemala (GT), Maryland (MD), Michigan (MI), Minnesota (MN), Mexico (MX), New Mexico (NM), Nevada (NV), Oregon (OR), Pennsylvania (PA), Texas (TX), Utah (UT), Venezuela (VE), Washingtzzson (WA), a combined Washington/Oregon (WA/OR) category, and Wisconsin (WI). Branch lengths represent genetic divergence, measured as the number of substitutions per site.

The common ancestor of sampled **C. posadasii** isolates was likely in Arizona, with a probability of 96%, while the common ancestor of sampled *C. immitis* isolates was likely in California, with a probability of 99%. However, the common ancestor of both sister species could not be determined with high probability, with estimates suggesting either California (30%) or Arizona (25%) as possible ancestral locations. The three *C. immitis* isolates from Utah, all originating from the same patient, appear to represent a travel-associated introduction from California, consistent with the patient’s documented travel history to San Diego and prior residence in Southern California.

## 3. Discussion

The genetic and evolutionary history of *C. immitis* and **C. posadasii** has been previously studied in endemic regions (e.g., Monroy-Nieto et al., 2023; Neafsey et al., 2010; Oltean et al., 2019); however, their presence in low-incidence states remains poorly characterized. Our study sought to bridge this gap by analyzing newly sequenced isolates from Utah, Colorado, and Nevada. Among the 27 newly sequenced clinical isolates, three were identified as *C. immitis* and 24 as **C. posadasii**. Utah was the only low-endemicity state where both species were detected, with **C. posadasii** identified in nine patients and *C. immitis* in a single individual. Although this case was likely travel associated, given the patient’s prior residence in California, the environmental presence of *C. immitis* in Utah has been previously documented. Soil samples collected from Dinosaur National Monument in 2006 confirmed the co-occurrence of both *C. immitis* and **C. posadasii**, providing evidence that both species can exist in the same geographic region (Johnson et al., 2014). In contrast, only **C. posadasii** was found among clinical isolates from Colorado and Nevada. Phylogenetic analyses revealed that isolates from the same state do not form monophyletic clades, suggesting multiple independent introductions rather than a single expansion. We identified at least seven introductions of **C. posadasii** into Utah, two into Nevada, and seven into Colorado.

Our findings align with previous studies that identified significant phylogeographic structure within *Coccidioides* species (Engelthaler et al., 2016; Monroy-Nieto et al., 2023). *Coccidioides posadasii* exhibits substantial population differentiation, forming distinct phylogeographic clades, including Arizona and Texas/Mexico/South America clades, as well as a recently identified Guatemala/Venezuela clade. In contrast, *C. immitis* populations appear geographically restricted primarily to California, with a distinct, recently recognized clade in Washington (Engelthaler et al., 2016; Monroy-Nieto et al., 2023). However, despite the apparent structure of these clades, we observed substantial genetic variation within each clade, likely reflecting large effective population sizes (Neafsey et al., 2010). Consequently, isolates did not cluster into strictly state-specific monophyletic clades. Our phylogenetic analyses revealed that **C. posadasii** predominates in low-endemicity regions, particularly Utah, Colorado, and Nevada. The presence of multiple independent introductions into these states suggests that *Coccidioides* migration is not a singular event but an ongoing process. These migration events likely stem from high-endemicity regions such as Arizona and Texas, where **C. posadasii** maintains greater genetic diversity. Our ancestral area reconstruction supports these findings, indicating that multiple invasions into low-endemicity areas occurred independently from distinct ancestral populations. The findings of this study have significant implications for public health.

Underdiagnosis and misdiagnosis of coccidioidomycosis remain persistent challenges, particularly in regions where the disease is not typically considered by clinicians (Eulálio et al., 2024; McHardy et al., 2023). Our results underscore the urgent need for continued surveillance and increased awareness among healthcare professionals and the public in low-incidence states. The broad geographic distribution of **C. posadasii** isolates further highlights the value of genomic surveillance in tracking pathogen spread, monitoring evolutionary dynamics, and identifying emerging epidemiological patterns.

Travel history is a critical component in understanding the transmission dynamics of coccidioidomycosis. Previous genomic studies have shown that many cases diagnosed in non-endemic regions are linked to travel to endemic areas, emphasizing that infections can be acquired far from where symptoms eventually appear (Monroy-Nieto et al., 2023; Qazi & Hancock, 2025). Our findings are consistent with this observation—for example, one patient’s *C. immitis* isolate was likely acquired in California, despite diagnosis occurring elsewhere. This supports the need for clinicians to consider travel exposure during diagnosis. Moreover, the ability of genomic epidemiology to trace isolates back to their probable geographic origins demonstrates the power of integrating molecular surveillance with patient travel histories to improve diagnostic precision and guide public health interventions (Barker et al., 2019; Monroy-Nieto et al., 2023). Strengthening these efforts is essential for tracking the spread of *Coccidioides* and anticipating shifts in disease distribution.

While our findings offer valuable insights, there are some limitations to consider. The genomic sampling strategy provides only a minimum estimate of the number of introductions into each location, as undetected lineages and uneven sampling across states may obscure the full extent of *Coccidioides* diversity and migration. Second, by sampling clinical isolates sent to a national diagnostic laboratory, we may exclude *Coccidioides* lineages that are associated with less severe symptoms or lineages that exist in enzootic cycles but are less infectious to humans (Taylor & Barker, 2019). Third, we were only able to conduct chart reviews for patients who were part of the University of Utah Health system, due to limits on protected health information associated with samples diagnosed at ARUP from other healthcare systems, diagnostic laboratories, and hospitals. Travel-associated infections further complicate interpretation, as some cases may reflect exposure in endemic regions rather than local transmission (Monroy-Nieto et al., 2023). Future research should expand genomic sampling across broader geographic and environmental ranges and collect detailed travel histories to clarify dispersal patterns. Long-term studies are also needed to track *Coccidioides* evolution and range expansion over time.

Our study extends previous work by characterizing the genetic diversity and migration history of *Coccidioides* in underrepresented regions. We find that **C. posadasii** has independently colonized states with low incidence of Valley fever multiple times, consistent with a pattern of ongoing gene flow, rather than a single introduction. These findings emphasize the importance of ongoing genomic surveillance to monitor ongoing *Coccidioides* dispersal.

## 4. Materials and Methods

### 4.1. Ethics Statement

This study was approved by the University of Utah Institutional Review Board (IRB_00164551). Consent procedures were conducted in accordance with the protocol approved by the IRB.

### 4.2. Sample Collection

We prospectively collected *Coccidioides-positive* isolates submitted to ARUP Laboratories, a national diagnostic laboratory from January 2023 to November 2024. We selected isolates from low-endemicity states, defined as those reporting fewer than 500 cases per year, that had limited available *Coccidioides* genomes, including 16 from Utah, 7 from Colorado, and 4 from Nevada (Fig 1). We additionally included prospectively sampled isolates from California due to its well-documented high endemicity (CDC; https://www.cdc.gov/valley-fever/hcp/clinical-overview/<u>\</u>) for *C. immitis* and relative lack of available whole genome sequences. We additionally included 136 sequences from previously published studies to achieve broad genomic representation across diverse geographic regions (Fig 1).

### 4.3. Whole Genome Sequencing

Genomic DNA was extracted from fungal cultures at ARUP using a Maxwell RSC Cell DNA purification kit (AS1370). DNA samples (1 to 25 ng) were enzymatically fragmented and libraries were prepared using the New England Biolabs NEBNext Ultra II FS DNA Library Prep kit (cat#E7805L) with an average insert size of 350 bp. PCR-amplified libraries were qualified on an Agilent Technologies 4150 TapeStation using a D1000 ScreenTape assay (cat# 5067-5582 and 5067-5583) and the molarity of adapter-modified molecules was defined by quantitative PCR using the Kapa Biosystems Kapa Library Quant Kit (cat#KK4824). Libraries were normalized and pooled in preparation for Illumina sequencing. We conducted paired-end whole genome sequencing (150 x 2 bp) on an Illumina NovaSeq X Series at the University of Utah High-Throughput Sequencing Core.

### 4.4. Variant Calling

Variant calling was performed using cocci-call (Marchetti et al., 2025), a bioinformatics pipeline specifically developed for species identification and detection of single nucleotide polymorphisms (SNPs) and other genomic variants in *Coccidioides*. Within cocci-call, raw sequencing reads were preprocessed by trimming low-quality bases (Phred < 20) and removing adapters using Trim Galore (stringency = 1) (https://github.com/FelixKrueger/TrimGalore). Quality control reports were generated with FastQC (https://www.bioinformatics.babraham.ac.uk/projects/fastqc/). To minimize false variant detection due to contamination, Kraken2 (Wood et al., 2019)was used to classify reads taxonomically, removing non-*Coccidioides* reads. Species assignment was based on the log2-transformed ratio of unique minimizers assigned to *C. immitis* or **C. posadasii**. Following this, high-quality reads were mapped to species-specific reference genomes (*C. immitis* GCF_000149335.2 and **C. posadasii** GCA_018416015.2) using BWA-MEM (v0.7.17) (Li & Durbin, 2009), and duplicate reads were marked with GATK 4.3 (McKenna et al., 2010). If species assignment remained ambiguous, reads were mapped to both references, with assignment determined by the highest mapping percentage. To minimize the impact of repetitive elements on our dataset, repetitive sequences were identified and removed using both NUCmer (Kurtz et al., 2004) and RepeatMasker (https://www.repeatmasker.org/). Variants were called using GATK HaplotypeCaller and GenotypeGVCFs, with sample ploidy set to 1. After variant calling, samples with a mean coverage lower than 30× were removed. Since the coalescent model assumes neutrality, we extracted only intergenic regions from the VCF using annotation data, processed with SnpEff (Cingolani et al., 2012). Finally, VCF files were converted to FASTA format using vcf2msa (available at: https://github.com/mmarchetti90/vcf2msa), retaining only variant sites.

### 4.5. Phylogenetic Analysis

We employed a maximum likelihood approach to reconstruct the relationship of the sampled isolates using IQ-TREE 2 (Minh et al., 2020). The multiple sequence alignment file was generated using cocci-call, including only variant sites from intergenic regions. Model selection was performed with ModelFinder Plus (Kalyaanamoorthy et al., 2017), which evaluates multiple substitution models based on the Bayesian Information Criterion and incorporates ascertainment bias correction to account for the absence of invariant sites. To ensure robust statistical support for the tree topology, we assessed branch support using 1000 replicates of ultrafast bootstrap approximation (Hoang et al., 2018). The phylogenetic tree was rooted using the midpoint rooting method, which places the root at the midpoint of the longest path between any two taxa, providing an optimal approximation when an outgroup is not available.

### 4.6. Ancestral Area Reconstruction

To infer the historical migration of *Coccidioides*, we used ancestral area reconstruction with the R package *ape* (Paradis et al., 2004). Specifically, we utilized the function ace to estimate ancestral character states and their associated uncertainty. We considered locality as a discrete character, which we defined as the state of the submitting diagnostic lab for newly sequenced isolates from ARUP and the US state or country of origin for previously published sequence data. In total, we included 18 localities: 15 US states and 3 countries. We reconstructed the most probable historical dispersal routes and potential origins of specific lineages.

### 4.7. Exposure histories

For patients who are diagnosed in Utah, we conducted retrospective chart reviews to collect information on symptoms, diagnosis, and exposure history, including any information recorded on possible occupational, recreational or travel exposures. We additionally mailed letters to patients who tested positive for coccidioidomycosis in Utah, inviting them to participate in the study, with a link to a RedCap survey, which included information on their exposure history and clinical symptoms. Patients/participants of any age were recruited.

## Data Availability

The authors confirm that all data underlying the findings are fully available without restriction. The FASTA file used for phylogenetic analyses is publicly available at https://github.com/emanuelmfonseca/coccidioides-phylogenetic-data.

